# PRAGMATIC - PRostate cancer diAGnosis and MAnagement – Triage In the Clinical care pathway

**DOI:** 10.1101/2024.03.05.24303844

**Authors:** Abishek Sharma, Teresa Campbell, Anthony Bates, Rincy John, Charlotte Adams, Aisling Brassill, Bryony Lennon, Philip Camilleri, Ami Sabharwal, Philip Charlton, Gerard Andrade, Mark Tuthill, Andrew Protheroe, Alastair D Lamb, Tom Leslie, Aaron Leiblich, Francisco Lopez, Clare Verrill, Fergus Gleeson, Ruth MacPherson, Freddie C Hamdy, Richard Bell, Richard J Bryant

## Abstract

**Background:** It is important to investigate, diagnose and commence treatment for locally advanced and metastatic prostate cancer quickly to optimise treatment outcomes. Since the introduction of national 2-week wait and 31/62-day targets in the United Kingdom for investigation of suspected prostate cancer over 2 decades ago, the clinical pathway has become increasingly complex. This may lead to some patients with the most clinically significant disease having the rapidity of their diagnosis and commencement of treatment compromised by resource use in diagnosing less significant, or clinically insignificant, disease.

**Methods:** We will conduct a retrospective review of timelines for diagnosis and commencement of treatment for all men referred to a tertiary unit for investigation of suspected prostate cancer on the 2-week wait pathway in a 3-month period in 2023. In parallel, we will introduce triaging of all new 2-week wait referrals in a prospective 3-month period, with a dedicated nurse navigator streamlining patients for the most rapid investigation and treatment, based on pre-specified risk criteria including PSA, pre-biopsy mpMRI findings including TNM staging, and histology results. We hypothesise that this bespoke triaging system, above and beyond the 2-week wait and 2022 Faster Diagnostic Pathway guidance issued by NHS England, will improve timings for investigation and commencement of treatment for the most clinically significant prostate cancer cases.

**Conclusions:** The use of in-house criteria for triaging and stratification of the most clinically urgent and significant prostate cancer cases, identified by a nurse specialist navigator, may improve clinical outcomes for patients with greatest need for rapid prostate cancer imaging, diagnosis and treatment.

## Introduction

Prostate cancer is the commonest malignancy diagnosed in men in the United Kingdom, with around 52,300 new diagnosed cases, and 12,000 disease specific deaths, annually ^1^. In 2022 in England and Wales 19% of patients presented with metastatic disease ^2^. Two-week wait referral guidelines for referral from primary to specialist care for investigation of suspected prostate cancer, and 31- and 62-day targets for diagnosis and treatment respectively, were introduced in 2000 as part of the National Health Service (NHS) cancer plan for England ^3^, with the aim of improving the survival of patients with cancer. More recently, FASTER diagnosis targets were introduced in the UK in 2019 as part of the NHS Long Term Plan aim to diagnose, or exclude, prostate cancer in all patients referred from primary to specialist care within 28 days, and to commence treatment within 31 days of diagnosis ^4^. However, whilst these laudable arbitrary targets may have led to improvements in the time-to-diagnosis and time-to-treatment of cancer, there is no evidence that they have led to improvements in mortality ^5^. The clinical pathway for investigation of suspected prostate cancer has become increasingly complex over the last decade, with introduction of pre-biopsy multiparametric magnetic resonance imaging (mpMRI) ^6,7^, the possibility to avoid biopsy based on mpMRI findings ^6,7,8^, cognitive or fusion targeted biopsy ^9^, either via the transrectal (local anaesthetic transrectal biopsy, TRUS) or local anaesthetic transperineal (LATP) biopsy routes, and use of novel molecular state-of-the-art imaging modalities for staging ^10^. Moreover, management options have become increasingly complex for localised disease, and may include active surveillance, radical surgery, radical radiotherapy with neoadjuvant androgen deprivation therapy (ADT), brachytherapy, and focal therapy ^7^. The increased complexity of the clinical pathway, coupled with an absence of risk-stratification within two-week wait referral criteria, and the need for patients to receive appropriate counselling of their options at each step of the pathway, has led to a risk that patients with the most clinically significant disease (i.e. de *novo* locally advanced or metastatic cases) may have the rapidity of their diagnosis and commencement of treatment compromised by intensive resource use in diagnosing the large pool of less significant, or clinically insignificant, disease cases.

In the case of localised (i.e. organ confined) prostate cancer, the two-week wait, and 31/62-day, or FASTER 28/62-day targets, are arbitrary objectives, and achieving these targets may not improve clinical outcomes. Recent evidence from the ProtecT trial in the UK demonstrates remarkably low clinical progression and mortality rates for clinically localised prostate cancer at a median of 15 years follow-up, regardless of whether patients were randomised to receive active monitoring, radical surgery or radical radiotherapy ^11^. During the recent COVID pandemic, clinical priority for treatment of urological conditions was temporarily diverted away from prostate cancer treatment, and instead was focussed on other urological malignancies such as bladder and renal cancer, along with time-critical benign conditions such as ureteric stone disease ^12^. Ironically the COVID pandemic required the urological community, and policymakers such as the British Association of Urological Surgeons (BAUS), to bring into sharp focus the need to prioritise treatment of those urological conditions most in need of rapid diagnosis and treatment. During the subsequent move back towards standard clinical practice following the covid pandemic, focus has moved back towards the 2-week wait and 28/62-day FASTER pathway, such as the recent description of a RAPID prostate cancer diagnosis pathway ^13^. However, whilst the aims of rapidly diagnosing all cases of prostate cancer may be laudable, this approach may not accelerate the delivery of care to those most in need and may inadvertently divert resources away from other time-critical urological conditions ^14^.

Clinicians recognise that high-risk localised, locally advanced, and metastatic prostate cancer require expedient investigation and commencement of treatment. However, it is pragmatic that lower-risk localised cases do not require rapid diagnosis or treatment. Indeed, very low-risk prostate cancer may not need diagnosis at all, with active surveillance being recommended, which may continue for life. At the present time the current 2-week wait, and 28/62-day, pathway targets and referral criteria are a ‘one size fits all’ approach to the targets for clinical assessment, investigation, and treatment of suspected prostate cancer, or for confirmed exclusion of the presence of this malignancy (**Figure 1**). The targets do not set out criteria with which to prioritise the potential ‘higher risk’ cases at referral (for example those at highest risk of having locally advanced or metastatic disease, such as those with a PSA ≥50 ng/ml at baseline) from the very start of the clinical pathway, nor do they prioritise 2-week wait referral patients for the most rapid prioritisation of imaging, biopsy, completion staging imaging, and commencement of treatment.

**Figure 1:**
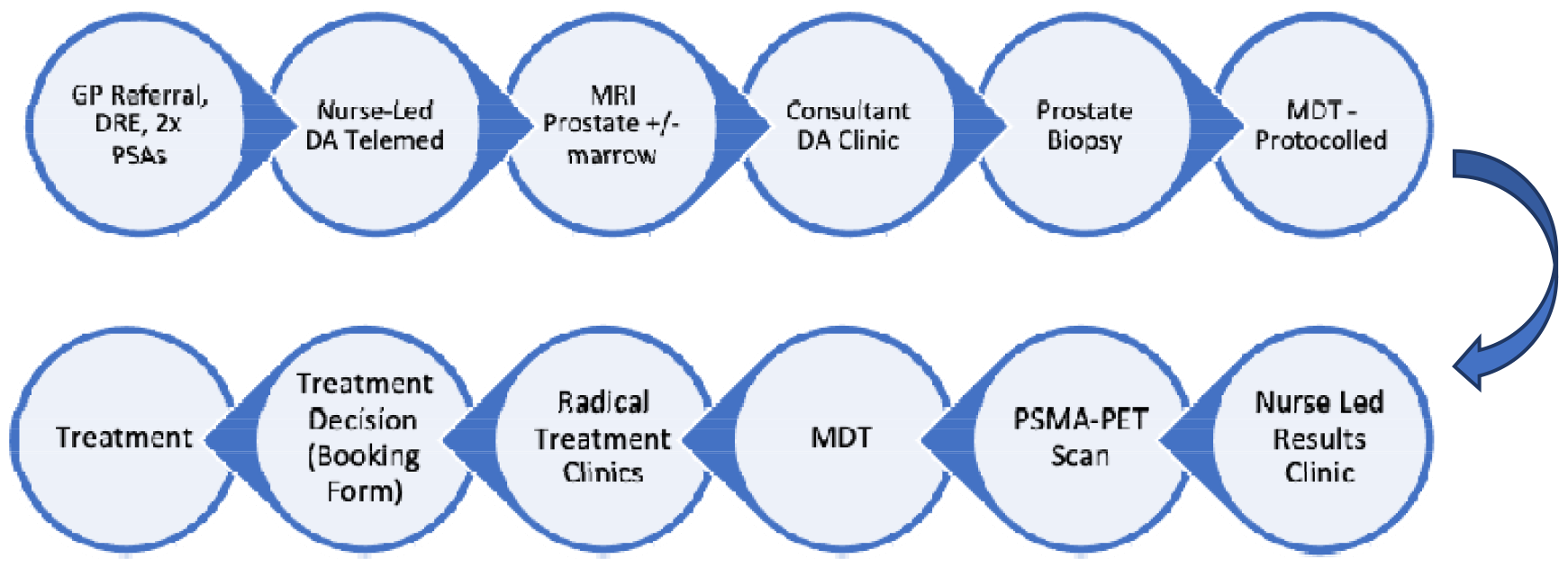
A flow chart indicative of the current patient pathway from primary care referral to commencement of treatment in Oxford. This flow chart gives the example of a patient with Gleason grade group 3 disease on biopsy, thus requiring a PSMA-PET CT scan for ‘completion staging’ based on our current local protocol.

There is an ongoing significant challenge and aspiration in the UK urological community to deliver a world class prostate cancer diagnosis and management pathway, whilst trying to meet 2-week wait targets, but that the ‘one size fits all’ approach is not meeting the urgent requirements of the highest risk cases of this malignancy.

## Methods

We have registered an actionable review of the entire 2-week wait and 28/62-day prostate cancer diagnosis and treatment pathway at our Institution (Oxford University Hospitals NHS Foundation Trust). We will undertake an internal Quality Improvement Project (QIP) to improve practice in this pathway for patients referred to our Institution with the highest need for rapid diagnosis and treatment. This process builds on ongoing work in the Urology Department to improve implementation of the arbitrary national pathway, such as ongoing evaluation of the pathway by Advanced Nurse Practitioners (ANPs) and clinical and non-clinical managers, recruitment of a ‘navigator’ to track and prioritise patients in the pathway based on need according to in-house criteria, and focussed help from the Cancer Improvement Team members and other stakeholders in the clinical pathway, including others aligned with the Urology Department (such as Radiology, Histopathology, and Clinical and Medical Oncology). This registered QIP (OUH reference number 8381) aims to allow all those engaged in the various multiple steps of the 2-week wait and 28/62-day pathway to triage the highest risk cases for the most rapid diagnosis and treatment, whilst simultaneously safeguarding against undue delay for lower-risk cases as an unintended consequence. This QIP has been reviewed by all stakeholders in the delivery of the clinical pathway at our Institution, including Urologists, ANPs, Cancer Specialist Nurses, Radiologists, Histopathologists, and Clinical and Medical Oncologists. This is important as some specialties involved in delivery of the pathway may need to slightly change practice to enable the proposed triaging protocol to be delivered effectively.

This QIP will involve a retrospective audit of all 2-week wait patients referred with suspected prostate cancer over a recent historical 3-month period, prior to the implementation of the triaging of the highest risk patient referrals. This will allow us to understand the timelines and bottlenecks in the pathway from initial referral to commencement of treatment, including the timeline for the highest risk patients, and those undergoing complex molecular imaging for triaging, which may add delay to the pathway. In addition to this retrospective evaluation of practice prior to the implementation of the internal triaging, we will prospectively audit a three-month period following implementation of the internal PRAGMATIC triaging, which will be based on previously published risk-stratification criteria such as the EAU risk groups for biochemical recurrence of localised and locally advanced prostate cancer ^7^ (Table 1). The pragmatic stratification process (Figure 2) will be dynamic and can be reviewed at every investigation step in the pathway, including at the Multi-Disciplinary Team (MDT) review. Patients may move upwards or downwards into a different stream, dependent upon the outcome of investigations at each node in the pathway. Patients will be assigned a triage stream based on their highest risk factor. For example, an individual with a PSA ≥50 ng/ml potentially has locally advanced or metastatic prostate cancer unless proven otherwise and will therefore automatically be placed in the highest risk category. As a further example, a patient with T3a prostate cancer on pre-biopsy mpMRI but Gleason grade group 1 on biopsy will likely have been under-sampled, thus requiring early and urgent review and likely repeat biopsy sampling. Certain streams of patients, such as ‘Yellow Stream’ patients, may require discussion in the prostate cancer MDT before they can be ‘stepped down’ and taken off the 2-week wait pathway. As an example of this, the NICE Prostate Cancer Guidelines in the UK state that patients with PIRADs 3-5 lesion on pre-biopsy mpMRI, but with negative prostate biopsy histology findings, require formal discussion in the prostate cancer MDT. We recognise, however, that there may also be a need for protocolised PRAGMATIC down-grading criteria to avoid potentially overburdening the highest risk patient streams. Our colour-coded PRAGMATIC ‘stratification streaming’ criteria are shown in **Table 1** and **Figure 2**. The ‘stratification streaming label’ will follow the patient from entry into the pathway until either commencement of treatment or discharge form the pathway. This will hopefully allow efficient triage of pre-biopsy mpMRI scans, prostate biopsy appointment, PSMA PET CT scan and other staging investigations, nurse led results clinics and consultant-led treatment option clinics, and dates for commencement of treatment. The over-arching aim is to prioritise a rapid pathway for the highest-risk patient groups, whilst simultaneously ensuring no patients are disadvantaged by this internal triaging process.

**Table 1:**
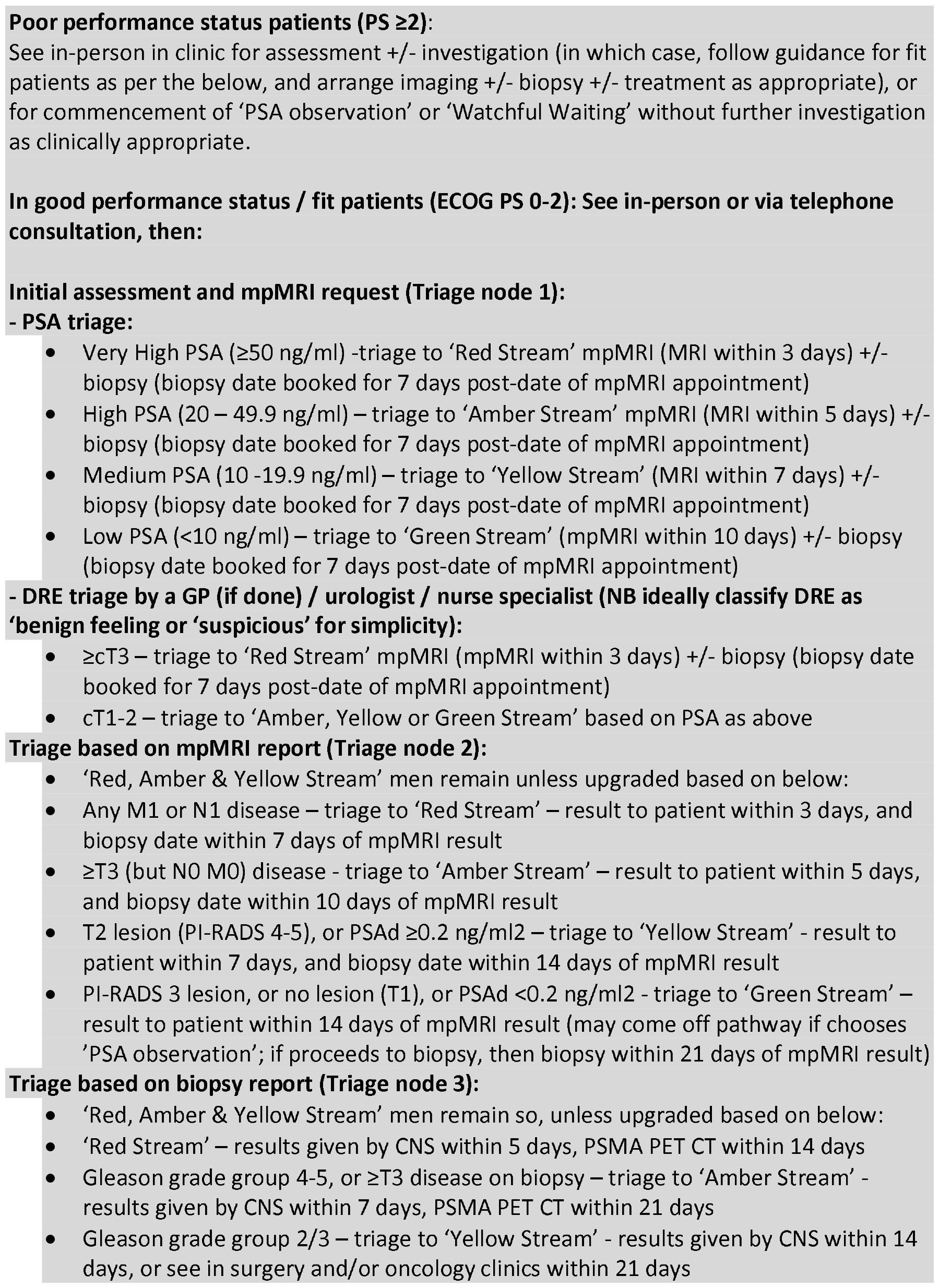
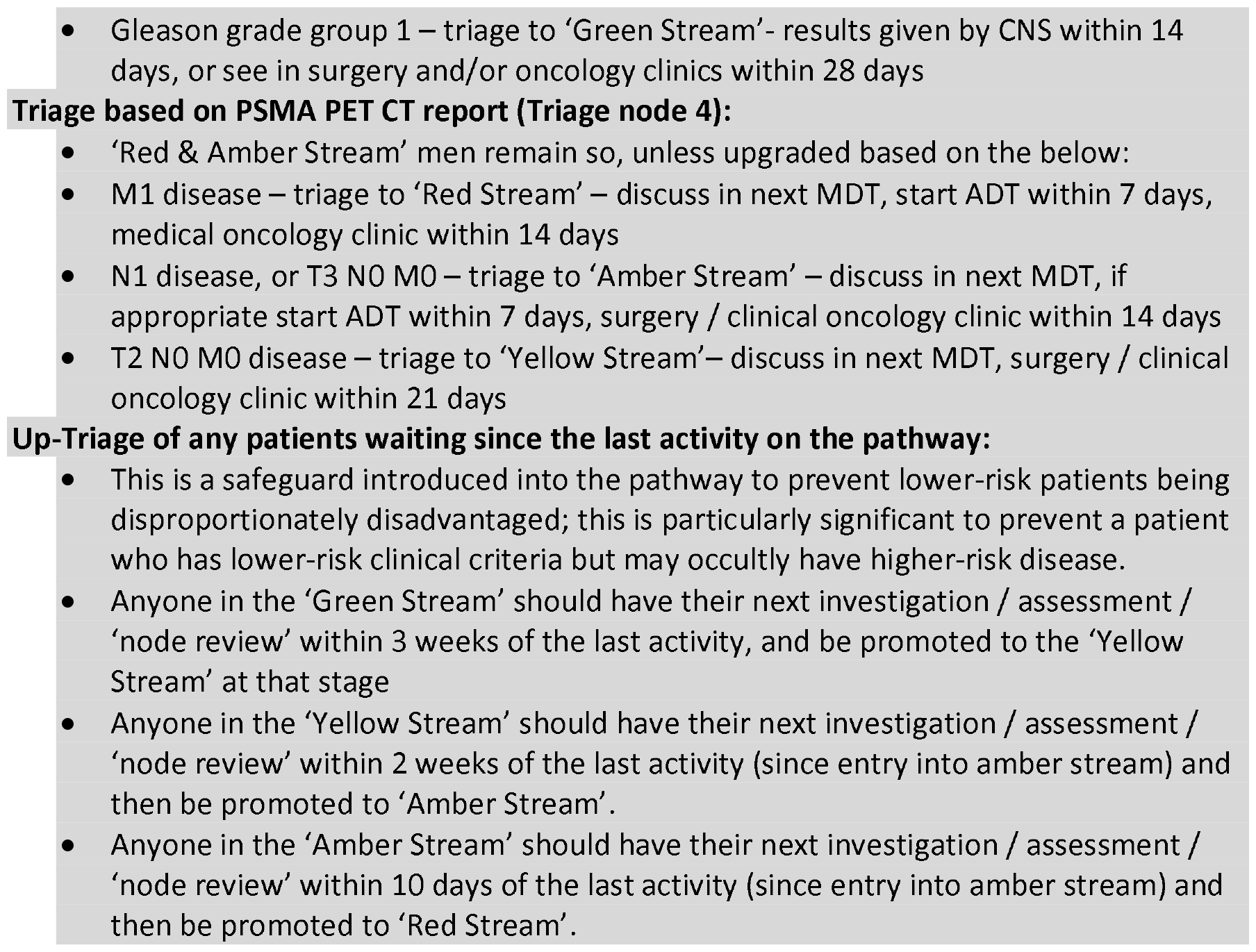
Indicative example of PRAGMATIC ‘stratification streaming’ criteria.

**Figure 2:**
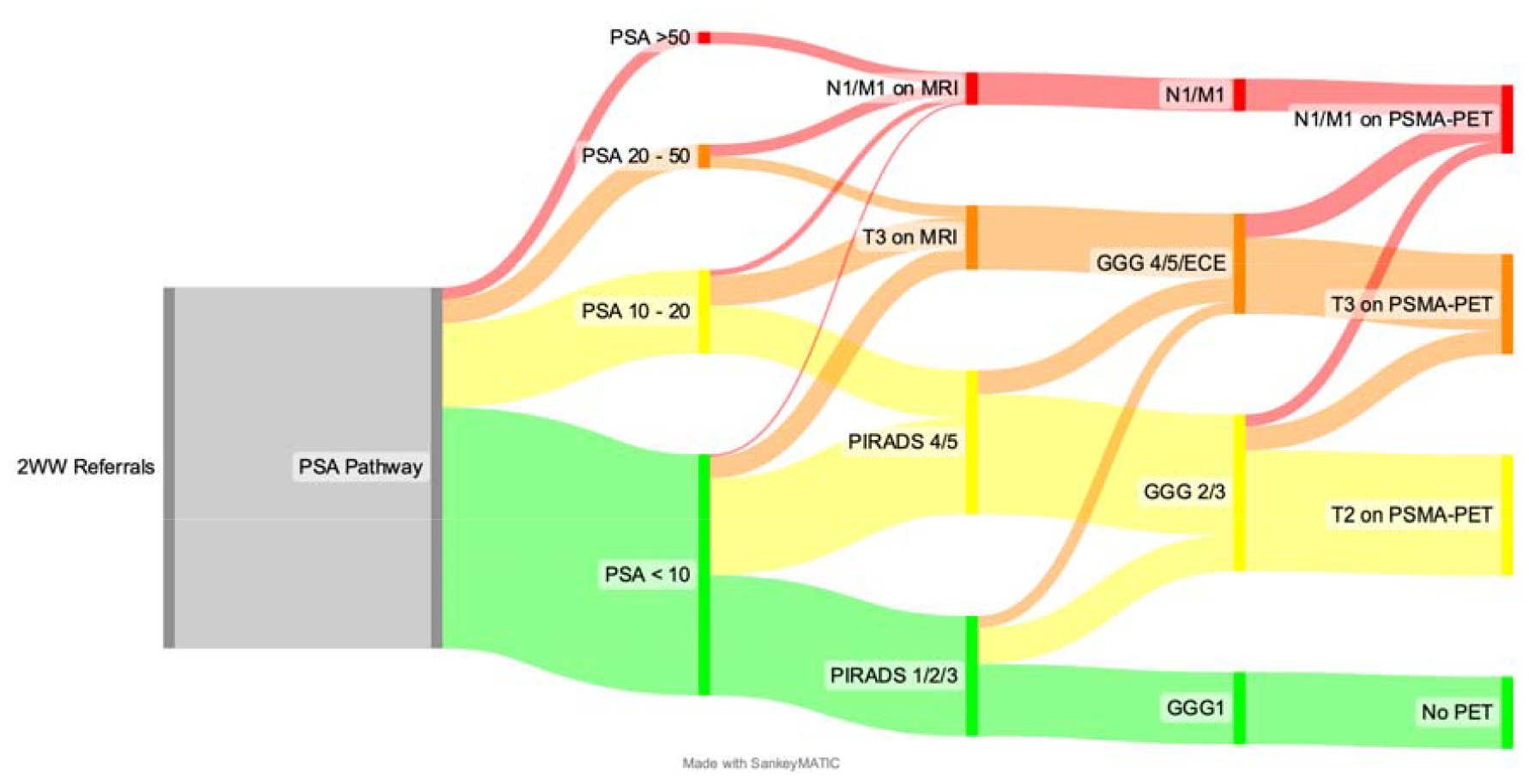
Example of the PRAGMATIC streaming criteria used in the 2-week wait Suspected Prostate Cancer Pathway. This triaging is for illustrative purposes only, and will be shaped by previously published risk categories, and by the results of the embedded internal audit of 6- months of patients referred on the 2-week wait Suspected Prostate Cancer Pathway (July-December 2022). The pathway will also include the removal of men after exclusion of prostate cancer, or who perhaps request ‘PSA observation’ in the context of a negative pre-biopsy MRI and low-risk of significant disease, or who are informed they have low-volume low- or favourable intermediate-risk prostate cancer suitable for Active Surveillance.

## Data Availability

All data produced in the present work are contained in the manuscript.

## Abbreviations

ADT: Androgen Deprivation Therapy
ANP: Advanced Nurse Practitioner
BAUS: British Association of Urological Surgeons
CNS: Clinical Nurse Specialist
DRE: Digital rectal Examination
EAU: European Association of Urology
ECOG: Eastern Cooperative Oncology Group
LATP: Local Anaesthetic TransPerineal
MDT: MultiDisciplinary Team
mpMRI: Multi-parametric Magnetic Resonance Imaging
NHS: National Health Service
NICE: National Institute for health and Care Excellence
PI-RADS: Prostate Imaging Reporting And Data System
PRAGMATIC: PRostate cancer diAGnosis and MAnagement – Triage In Clinical care pathway.
PSA: Prostate Specific Antigen
PSAd: Prostate Specific Antigen density
PSMA PET CT: Prostate Specific Membrane Antigen Positron Emission Tomography– Computed Tomography
QIP: Quality Improvement Project
TNM: Tumour, Nodes, Metastasis
TRUS: TransRectal Ultrasound

## References

1. Prostate Cancer Statistics, Cancer Research UK. Available at: https://www.cancerresearchuk.org/health-professional/cancer-statistics/statistics-by-cancer-type/prostate-cancer#heading-One Accessed 05/02/2024

2. National Prostate Cancer Audit. Available at: https://www.npca.org.uk/content/uploads/2024/01/NPCA-SotN-infographic.pdf Accessed 05/02/2024

3. Department of Health. The NHS cancer plan. Department of Health, London. 2000

4. The NHS Long Term Plan. NHS England, 2019. Available at: https://www.longtermplan.nhs.uk/

5. Oke JL, Brown SJ, Senger C, and Gilbert Welch H. Deceptive measures of progress in the NHS long-term plan for cancer: case-based vs. population-based measures. Br J Cancer 2023; 129(1) :3–7.

6. Kasivisvanathan V, Rannikko AS, Borghi M, Panebianco V, Mynderse LA, Vaarala MH et al. MRI-Targeted or Standard Biopsy for Prostate-Cancer Diagnosis. N Engl J Med 2018; 378(19): 1767–1777.

7. Mottet N, van den Bergh RCN, Briers E, Van den Broeck T, Cumberbatch MG, De Santis M et al. EAU-EANM-ESTRO-ESUR-SIOG Guidelines on Prostate Cancer-2020 Update. Part 1: Screening, Diagnosis, and Local Treatment with Curative Intent. Eur Urol 2021; 79(2): 243–262.

8. Schoots IG and Padhani AR. Risk-adapted biopsy decision based on prostate magnetic resonance imaging and prostate-specific antigen density for enhanced biopsy avoidance in first prostate cancer diagnostic evaluation. BJUI 2023; 127: 175–178.

9. Omer A and Lamb AD. Optimizing prostate biopsy techniques. Curr Opin Urol 2019; 29(6): 578–586.

10. Hofman MS, Lawrentschuk N, Francis RJ, Tang C, Vela I, Thomas P, et al. Prostate-specific membrane antigen PET-CT in patients with high-risk prostate cancer before curative-intent surgery or radiotherapy (proPSMA): a prospective, randomised, multicentre study. Lancet 2020; 395(10231): 1208–1216.

11. Hamdy FC, Donovan JL, Lane JA, Metcalfe C, Davis M, Turner EL et al. Fifteen-Year Outcomes after Monitoring, Surgery, or Radiotherapy for Prostate Cancer. N Engl J Med 2023; 388(17): 1547–1558.

12. BAUS, British Association of Urological Surgeons. COVID-19 strategy for the Interim management of prostate cancer prepared by the BAUS Section of Oncology. Version 1: 19 March 2020. Available from: https://www.cmcanceralliance.nhs.uk

13. Eldred-Evans D, Connor MJ, Bertoncelli Tanaka M, Bass E, Reddy D, Walters U, et al. The rapid assessment for prostate imaging and diagnosis (RAPID) prostate cancer diagnostic pathway. BJU Int 2023; 131(4): 461–470.

14. Lopez JF and Bryant RJ. The ‘Rapid Access Prostate Imaging and Diagnosis’ (RAPID) diagnostic pathway: what is the rush? BJU Int 2023; 131(4): 377–379.

